# A common *TMPRSS2* variant protects against severe COVID-19

**DOI:** 10.1101/2021.03.04.21252931

**Authors:** Alessia David, Nicholas Parkinson, Thomas P Peacock, Erola Pairo-Castineira, Tarun Khanna, Aurelie Cobat, Albert Tenesa, Vanessa Sancho-Shimizu, GenOMICC Investigators, ISARIC4C Investigators, Jean-Laurent Casanova, Laurent Abel, Wendy S. Barclay, J. Kenneth Baillie, Michael JE Sternberg

**Author notes:** **Corresponding Author:** Dr Alessia David MD, MSc, PhD, Centre for Integrative System Biology and Bioinformatics, Department of Life Sciences, Imperial College London, London, UK.

## Abstract

Infection with SARS-CoV-2 has a wide range of clinical presentations, from asymptomatic to life-threatening. Old age is the strongest factor associated with increased COVID19-related mortality, followed by sex and pre-existing conditions. The importance of genetic and immunological factors on COVID19 outcome is also starting to emerge, as demonstrated by population studies and the discovery of damaging variants in genes controlling type I IFN immunity and of autoantibodies that neutralize type I IFNs. The human protein transmembrane protease serine type 2 (TMPRSS2) plays a key role in SARS-CoV-2 infection, as it is required to activate the virus’ spike protein, facilitating entry into target cells. We focused on the only common *TMPRSS2* non-synonymous variant predicted to be damaging (rs12329760), which has a minor allele frequency of ∼25% in the population. In a large population of SARS-CoV-2 positive patients, we show that this variant is associated with a reduced likelihood of developing severe COVID19 (OR 0.87, 95%CI:0.79-0.97, p=0.01). This association was stronger in homozygous individuals when compared to the general population (OR 0.65, 95%CI:0.50-0.84, p=1.3×10^−3^). We demonstrate *in vitro* that this variant, which causes the amino acid substitution valine to methionine, impacts the catalytic activity of TMPRSS2 and is less able to support SARS-CoV-2 spike-mediated entry into cells.

*TMPRSS2* rs12329760 is a common variant associated with a significantly decreased risk of severe COVID19. Further studies are needed to assess the expression of the *TMPRSS2* across different age groups. Moreover, our results identify TMPRSS2 as a promising drug target, with a potential role for camostat mesilate, a drug approved for the treatment of chronic pancreatitis and postoperative reflux esophagitis, in the treatment of COVID19. Clinical trials are needed to confirm this.

## Main

The severe acute respiratory syndrome like coronavirus (SARS-CoV-2) has infected over 107 million individuals globally and has caused more than 2.3 million deaths. SARS-CoV-2 infection has a broad clinical spectrum, ranging from asymptomatic or mild symptomatic, to a life-threatening presentation requiring admission to intensive care. Age, and to a much lesser extent male gender and underlying clinical conditions, such as cardiovascular disease, obesity and diabetes, are known risk factors associated with an increased COVID19 morbidity and mortality. ^1,2^ The role of an individual’s genetic background has recently emerged as an additional, yet not clearly understood, risk factor for COVID19 ^3,4,5^. Rare genetic variants in genes involved in the regulation of type I interferon (IFN) immunity, including autosomal recessive IRF7 and IFNAR1 deficiencies, have been identified in patients with life-threatening COVID19 ^5^. Autoantibodies to type I IFNs also account for at least 10% of cases of critical COVID19 pneumonia ^6^. Moreover, genome-wide association studies (GWAS) have discovered genetic haplotypes spanning several genes that are associated with COVID19 severity ^2,7,3^.

The transmembrane protease serine type 2 (TMPRSS2) protein has a key role in coronavirus infections, including SARS-CoV-2, as it is required for priming the virus’ spike (S) glycoprotein through its cleavage, thus facilitating endosome-independent entry into target cells ^8,9^. *TMPRSS2*, which is part of the type 2 transmembrane serine proteases (TTSPs) family, is characterized by androgen receptor elements located upstream to its transcription site ^10^. As well as cleaving and activating viral glycoproteins of coronaviruses and influenza A and B viruses ^11^, TMPRSS2 is subjected to autocleavage, which results in the liberation of its soluble catalytic domain ^12^. The conditions under which autocleavage of TMPRSS2 and other members of the TTSPs family occurs are yet to be elucidated.

TMPRSS2 is expressed in lung and bronchial cells ^13^, but also in the colon, stomach, pancreas, salivary glands and numerous other tissues ^14^. Moreover, it is co-expressed in bronchial and lung cells with the angiotensin-converting enzyme 2 (ACE2) ^13^, which is the best described SARS-CoV-2 cellular receptor ^15^. In the olfactory epithelium of mice, the expression of *TMPRSS2*, but not *ACE2*, appears to be age-related and greater in old compared to young animals ^16^. Similarly, a recent study showed that expression of TMPRSS2 in mouse and human lung tissue is also age-related ^17^. Studies in *TMPRSS2* knock out (KO) mice reported reduced SARS-CoV and MERS-CoV replication in the lungs compared to wild-type mice, and a reduced proinflammatory viral response, especially cytokine and chemokine response via the Toll-like receptor 3 pathway ^18, 19^. We have recently shown that TMPRSS2 expression permits cell surface entry of SARS-CoV-2, allowing the virus to bypass potent endosomal restriction factors ^20^. *In vitro* studies have shown that TMPRSS2 inhibitors prevent primary airway cell and organoid infection by SARS-CoV and SARS-CoV-2 ^21, 20, 22^. In animal studies, mice infected with SARS-CoV and treated with the serine protease inhibitor camostat mesilate had a high survival rate ^23^. Recently, camostat mesilate (which, in Japan, is already approved for patients with chronic pancreatitis and postoperative reflux esophagitis) was shown to block SARS-CoV-2 lung cell infection *in vitro* ^8, 20^.

Based on the above data from animal models and cell-based studies supporting a protective role of a knock out *TMPRSS2* on coronavirus infection (including SARS and MERS), we hypothesized that naturally-occurring *TMPRSS2* genetic variants affecting the structure and function of the TMPRSS2 protein may modulate the severity of SARS-CoV-2 infection (here defined by the presence of respiratory symptoms severe enough to require a minimum of hospital admission).

We analysed 378 *TMPRSS2* genetic variants reported in GnomAD (v2.1.1), the database of population genetic variations. We studied the evolutionary conservation of TMPRSS2 amino acids and the impact of amino acid substitution on TMPRSS2 protein structure (described in Methods). As no experimental structure of TMPRSS2 is yet available, we generated a 3D structural model using homology modelling (Figure 1). We identified the chemical and physical bonds that stabilize the TMPRSS2 structure (i.e. hydrogen bonds, cysteine and salt bridges, as detailed in the Methods) and are affected by amino acid substitutions naturally occurring in the human population. A total of 137 variants were predicted damaging to the structure and/or function of TMPRSS2. Of these, 136 variants are extremely rare in the human population, with an average minor allele frequency (MAF) of 9.67×10^−6^ (cumulative MAF of 7.3×10^−4^) and are, therefore, unlikely to be of use as a marker of COVID19 infection severity in the general population. The remaining variant, rs12329760 C>T, is predicted damaging and causes the substitution of an evolutionary conserved valine to methionine (p.Val160Met on Ensembl transcript ENST00000332149.5 and Val197Met on the longer Ensembl transcript ENST00000398585.3). Overall, the minor allele frequency (MAF) of this variant is 0.25 in the human population, with 6.7% homozygous individuals (9,587 T/T homozygotes out of 141,456 individuals sequenced as part of the GnomAd project). Under Hardy-Weinberg equilibrium and MAF of 0.25, it is expected that 37% of individuals will be heterozygous for this variant. The MAF of rs12329760 T varies according to ethnicities and ranges from 0.14 in Ashkenazi Jewish to 0.38 in East Asian populations (0.15 in Latino, 0.23 in non-Finnish Europeans, 0.25 in South Asians, 0.29 in African/African Americans and 0.37 in Finnish Europeans). This highly conserved valine occurs in the scavenger receptor cysteine-rich (SRCR) domain, whose function within TMPRSS2 is still not fully understood, although a role in ligand and/or protein interaction has been proposed ^24^. Indeed, this domain is present in several proteins involved in host defence, such as CD5, CD6 and Complement factor I ^25, 26^.

**Figure 1.**
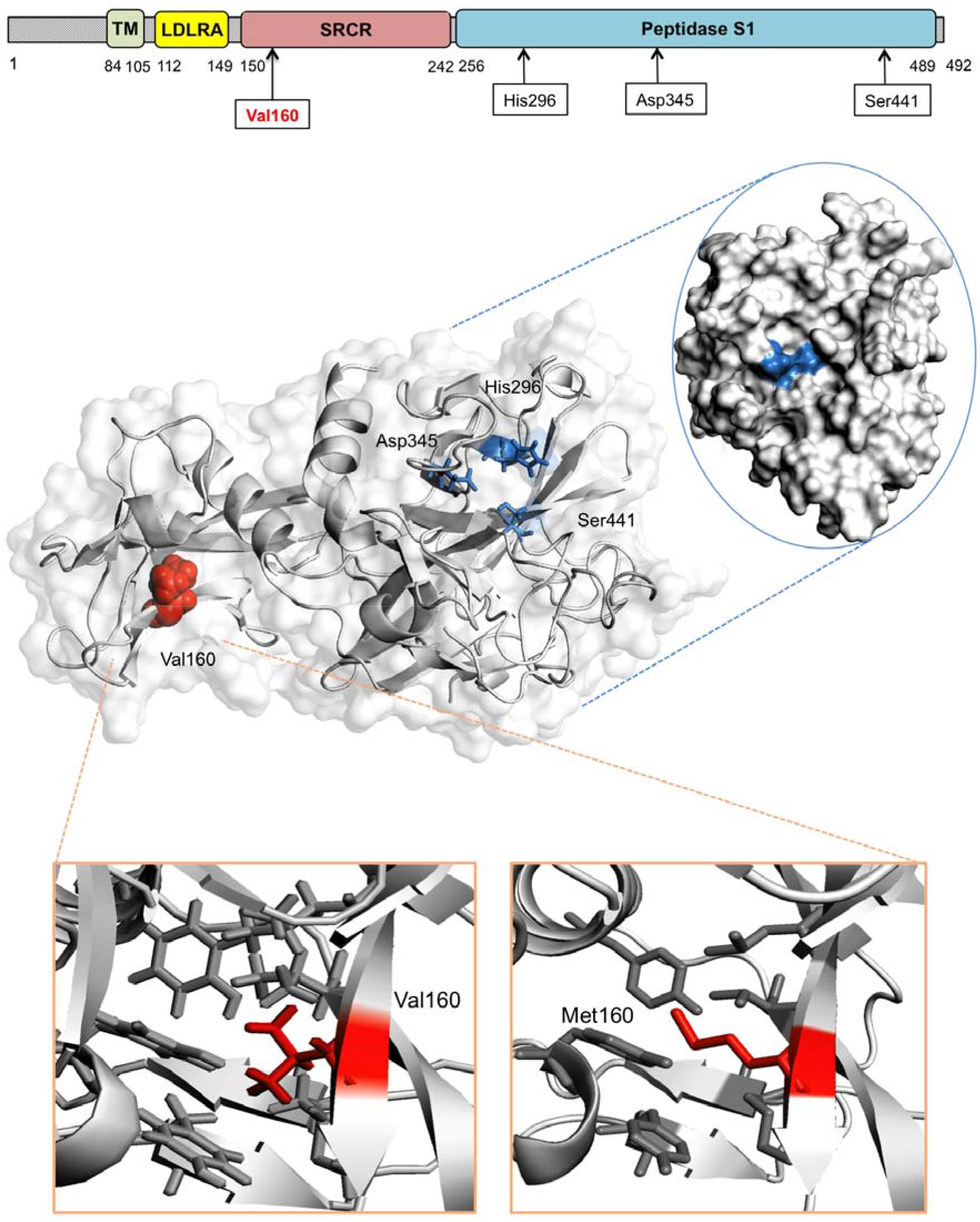
The TMPRSS2 protein and the p.Val160Met variant. The TMPRSS2 protein is composed of a cytoplasmic region (residues 1-84), a transmembrane region (TM, residues 85-105) and an extracellular region (residues 106-492). The latter is composed of three domains: the LDLR class A (residues 112-149), the scavenger receptor cysteine-rich domain (SRCR) (residues 150-242) and the Peptidase S1 (residues 256-489), which contains the protease active site: residues His296, Asp345 and Ser441. The 3D model of the extracellular region residues 145-491 corresponding to domains SRCR-2 (in green) and Peptidase S1 (in blue) is presented. Valine 160 (Val 160, depicted as a red sphere on the cartoon), which harbours variant p.Val160Met, occurs in the SRCR domain and spatially far from the TMPRSS2 catalytic site (mapped onto the surface of TMPRSS2). The transmembrane serine protease hepsin was used as a template to generate the model (PDB: 1Z8G, chain A, X-ray structure with 1.55Å resolution; model confidence 100%, sequence identity to target sequence= 35%).

We first analysed the relation between *TMPRSS2* rs12329760 and life-threatening SARS-CoV-2 infection in 2,244 critically-ill (defined in Methods) hospitalized COVID19-positive patients (table 1) recruited as part of the GenOMICC (genomicc.org) and ISARIC 4C (isaric4c.net) projects, and 11,220 ancestry-matched individuals without a COVID19-positive PCR test from the UK BioBank, who acted as controls. Under an additive model, we found that the minor T allele of rs12329760 was significantly associated with a protective effect against severe COVID19 in individuals of European ancestry (1,676 cases, 8,379 controls) with an OR of 0.87 (95%CI:0.79-0.97, p=0.01). A protective effect was also observed in individuals of East Asian ancestry (149 cases, 745 controls; OR 0.64, 95%CI:0.43-0.95, p=0.03). Similar effect sizes were observed in South Asians and Africans, but did not reach statistical significance, most likely as a result of the small sample size (Figure 2). We further confirmed this protective effect on a trans-ethnic meta-analysis, using the entire cohort of 2,244 patients (OR 0.84, 95%CI:0.77-0.93, p=5.8×10^−4^, Figure 2, panel A). A heterogeneity analysis suggested that the T allele has a similar effect across different ethnicities (p=0.47). To ascertain that this association was not an artefact due to population bias in the UK BioBank controls, the results from the European cohort were confirmed on an independent control population (45,875 unrelated individuals of European ancestry from 100K Genomes ^27^ : OR 0.89, 95%CI:0.81-0.99, p=0.02). Under a recessive model (i.e. individuals homozygous for the T allele), the trans-ethnic meta-analysis on 2,244 critically-ill COVID19 patients estimated an OR of 0.65 for TT homozygotes (95%CI:0.50-0.84, p=1.3×10^−3^). In subset analyses, the OR was estimated at 0.70 (95%CI:0.52-0.95, p=0.024) in Europeans, and 0.28 (95%CI:0.09-0.82, p=0.019) in East Asians versus their corresponding ancestry-matched controls (Figure 2, panel B).

**Table 1.** Patient characteristics.

|  | **GenOMICC/**<br>**ISARIC 4C** |
| --- | --- |
| Patients, n. | 2,244 |
| Females, n. (%) | 624 (30) |
| Age (years), mean±SD | 57.3 ±12.1 |
| Admitted to ITU/ICU, n. (%) | 1557 (74) |

**Figure 2.**
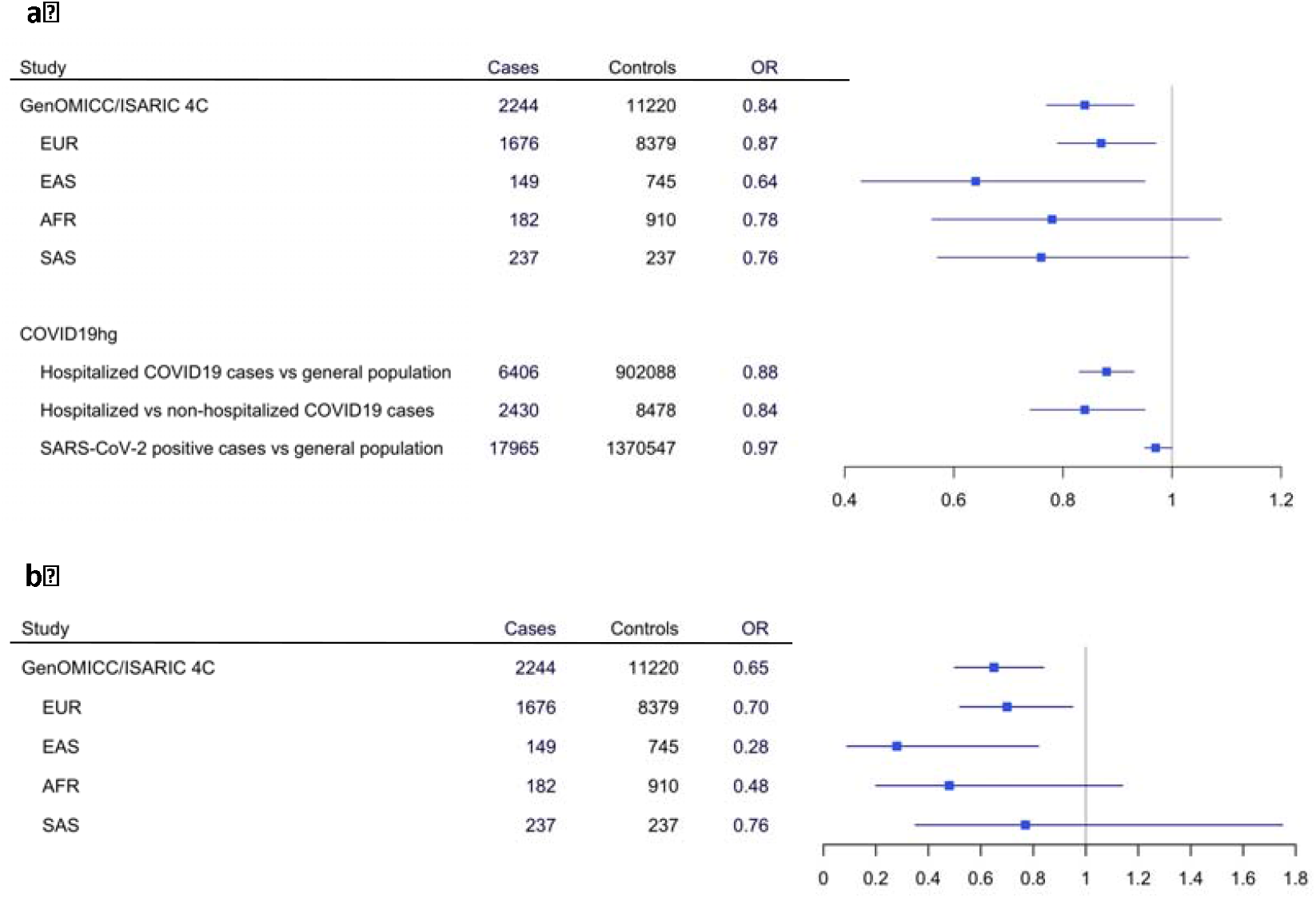
Association of *TMPRSS2* rs12329760 to COVID19 severity. Results are presented for the additive (a) and the recessive (b) model using different COVID19-positive patient cohorts. The results from large GWAS meta-analyses performed as part of the COVID19hg initiative (https://www.covid19hg.org/) are also shown.

In a second step, we investigated results from large GWAS meta-analyses performed in the context of the COVID19 Host Genetics Initiative2 (COVID19hg, available at https://www.covid19hg.org/, release January, 2021). Compared with the general population, the minor T allele of rs12329760 was significantly associated with a protective effect against severe COVID19 (patients requiring hospitalization for COVID19): 12,888 cases versus 1,295,966 individuals; OR 0.93, 95%CI:0.90-0.97, p=4.39×10^−**4**^. Furthermore, in a subgroup analysis, the rs12329760 had a protective effect against severe (SARS-CoV-2 confirmed infection requiring hospitalization, n=5,773) when compared to non-severe COVID19 (i.e. SARS-CoV-2 confirmed infection not requiring hospitalization 21 days after the test, n of infected patients=15,497): OR 0.92, 95%CI:0.87-0.98, p=2.25×10^−2^. Finally, there was no difference (p=0.57) in the prevalence of the T allele between the general population (n=1,668,938) and pooled individuals with a laboratory-confirmed SARS-CoV-2 infection (including hospitalized and life-threatening COVID19 cases from the metanalyses previously described) or with a self-reported or physician-confirmed COVID diagnosis (total n=36,590 cases).

Although an overlap in the control sets used in these meta-analysis may be present, these results are consistent with our hypothesis that the TMPRSS2 rs12329760 variant has a protective effect against severe and/or life-threatening COVID19. However, studies examining the prevalence of this variant in SARS-CoV-2 infected asymptomatic or pauci-symptomatic individuals are needed to ascertain its protective effect against mild viral infection.

The allele frequency of *TMPRSS2* rs12329760 (data from GnomAD population database) varies across different populations and is higher in East Asian and Finnish individuals (MAF 0.38 and 0.37, respectively) compared to south Asians (MAF 0.25) and Europeans (non-Finnish, MAF 0.23). The lowest frequency of the T allele is reported in Latino and Jewish-Ashkenazi individuals (MAF 0.15). Genotyping of the *TMPRSS2* rs12329760 variant on large COVID19 cohorts of patients of non-European genetic ancestry is needed to assess its role in determining the differences in the severity of COVID19 across various populations (e.g. between East Asia and Europe ^28^). Indeed, a recent study showed a lower T allele frequency in a small cohort of Chinese patients with life-threatening COVID19 compared to the population frequency ^29^. Although the differences in the proportion of SARS-CoV-2 patients who develop severe COVID19 across different populations ^28^ are more likely to be explained by social behaviour, public health measures to curb outbreaks, exposure to other viruses and immunological factors, human genetic variation across different populations may also marginally contribute to the observed differences.

To investigate the phenotypic effect of the TMPRSS2 V160M variant, we co-transfected 293Ts cells with ACE2 and either TMPRSS2 wild type (TMPRSS2_WT_) or V160M (TMPRSS2_V160M_), as previously described ^20^. We and others previously observed that co-expression of TMPRSS2 and ACE2 results in rapid cleavage of ACE2. We, therefore, used a mutant ACE2 that cannot be degraded by TMPRSS2 ^30^. Two additional TMPRSS2 variants were included as controls: the catalytically *inactive* S441A (TMPRSS2_S441A_) and the catalytically *active* R255Q (TMPRSS2_R255Q_), that is unable to autocleave ^12^. First, we investigated the autocleavage pattern of the different TMPRSS2 variants. The N-terminal membrane-bound part of TMPRSS2 can exist as different cleaved intermediates: a full-length uncleaved form of approximately 55 kDa, an intermediately cleaved form, and a fully cleaved form of 20 kDa. The latter is the product of TMPRSS2 autocleavage at arginine 255, which results in the release of the catalytically active protease domain in the extracellular space ^12^. Wild type TMPRSS2 is expressed as roughly equal amounts of full-length and fully cleaved forms, with a small amount of intermediately cleaved product. As expected, the catalytically inactive TMPRSS2_S441A_ and the non-autocleavable TMPRSS2_R255Q_ resulted in only the full-length TMPRSS2 being expressed. However, TMPRSS2_V160M_ resulted in a significantly higher proportion of full-length (55 kDa), and significantly lower proportion of fully cleaved protein (20 kDa) (p<0.05, Student’s t-test), suggesting that the V160M substitution exerts a partial inhibitory effect on the proteolytic autocleavage of TMPRSS2 (see Figure 3A-C).

**Figure 3.**
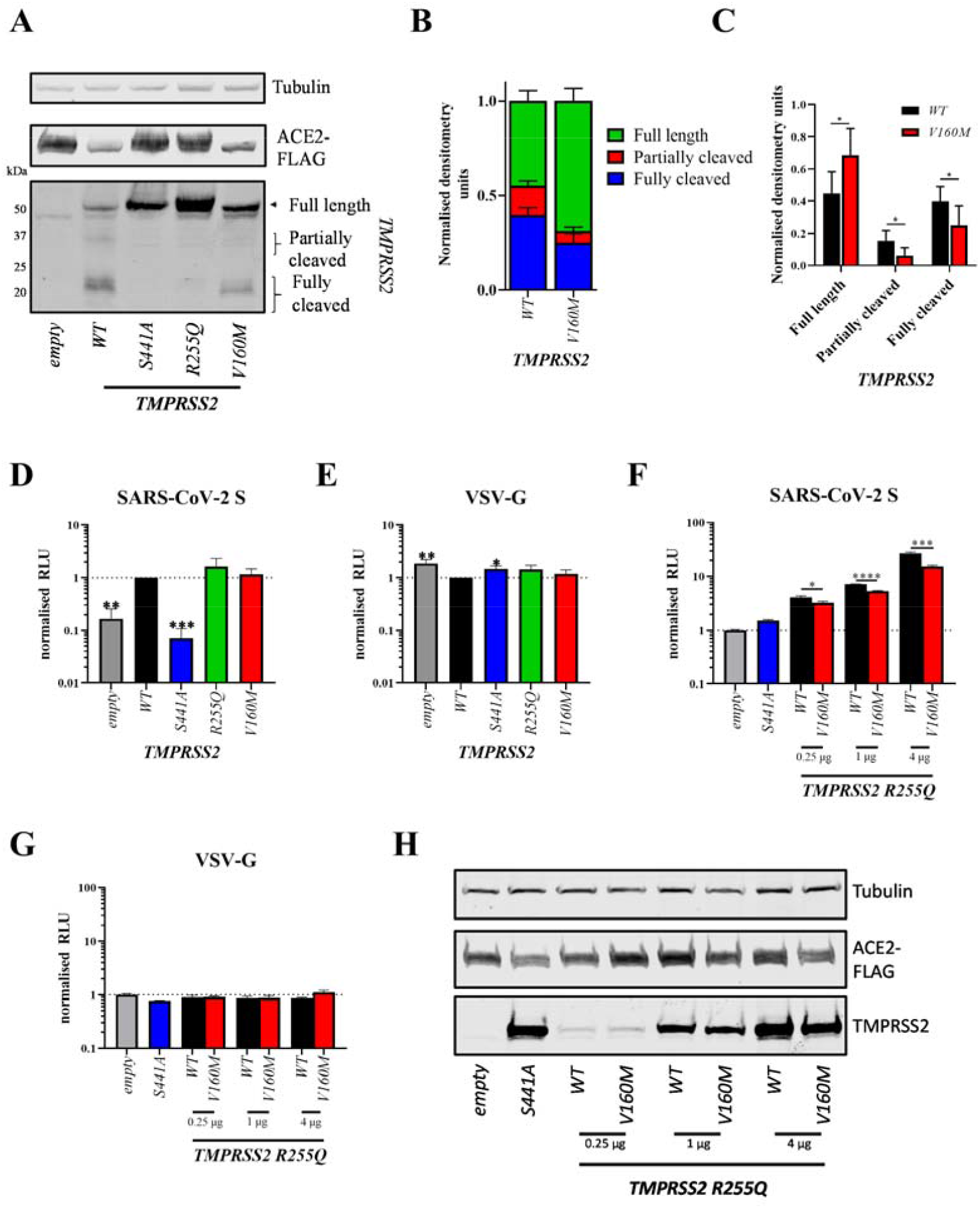
Phenotypic impact of the TMPRSS2 V160M variant on autocleavage and SARS-CoV-2 spike-mediated entry. (A) Western blot analysis of TMPRSS2 autocleavage after expression in HEK 293Ts. (B,C) densitometry was determined in ImageJ and shows mean±standard deviation from 6 independent repeats. Statistics determined by two-tailed Student’s t-test. Entry of lentiviral pseudotypes expressing (D, F) SARS-CoV-2 spike glycoprotein or (E, G) Vesicular stomatitis virus glycoprotein (VSV-G) into HEK 293Ts co-expressing ACE2-FLAG and either empty vector or TMPRSS2 variants. Data shows mean±standard deviation of 3 independent repeats from different weeks, normalised to WT TMPRSS2. (D, E) Statistics determined by one-way ANOVA on Log-transformed data (after determining log normality by the Shapiro-Wilk test and QQ plot). (F, G) statistics determined by two-tailed Student’s t-test. (H) Western blot analysis of TMPRSS2 autocleavage mutant (R255Q) titration with or without the V160M substitution. µg values indicate the amount of TMPRSS2 plasmid added to each condition. RLU, relative luminescence units. *, 0.05 ≥ P > 0.01; **, 0.01 ≥ P > 0.001, ***, 0.001 ≥ P

Subsequently, we investigated the effect of TMPRSS2_V160M_ on promoting viral entry, using a previously described SARS-CoV-2 pseudovirus entry assay ^20^. Pseudovirus expressing the glycoprotein from the vesicular stomatitis virus (VSV-G) was used as a control, as this virus enters cells in a TMPRSS2-independent manner ^20^. Briefly, cells co-transfected with ACE2 and TMPRSS2 wild type or variants were incubated with the pseudovirus (as described in ^20, 31^) and after 48h, luminescence was measured. TMPRSS2_WT_ enhanced viral entry by ∼5-fold compared to empty vector, while the catalytically dead TMPRSS2_S441A_ showed no enhancement (Figure 3D). The non-autocleavable mutant TMPRSS2_R255Q_ showed a similar enhancement, suggesting that autocleavage is dispensable for optimal TMPRSS2-mediated enhancement. TMPRSS2_V160M_ showed no significant difference in viral entry compared to the TMPRSS2_WT_. Overall, expression of catalytically active TMPRSS2 proteins slightly inhibited VSV-G mediated entry (Figure 3E).

The partial inhibitory effect exerted by the V160M variant on the proteolytic autocleavage of TMPRSS2 resulted in a far greater proportion of uncleaved, surface-expressed TMPRSS2_V160M_ compared to TMPRSS2_WT_. We, therefore, re-assessed whether TMPRSS2_V160M_ affects SARS-CoV-2 S-expressing pseudovirus entry by using the double mutant TMPRSS2_R255Q/V160M_ (which cannot autocleave) to control for protein cell-surface expression. Under these conditions, and across a range of plasmid concentrations, TMPRSS2_R255Q/V160M_ showed a significantly reduced ability to promote SARS-CoV-2 S-expressing pseudovirus compared to TMPRSS2_R255Q_ alone, despite equal protein expression (Figure 3F, H). Again, TMPRSS2_R255Q/V160M_ had no effect on VSV-G-mediated entry (Figure 3G).

Overall, our results suggest that the V160M substitution results in a moderately less catalytically active TMPRSS2, which is less able to autocleave and prime the SARS-CoV-2 spike protein. However, very little is known on TMPRSS2 and further extensive in vitro and in vivo studies on its pathophysiology are necessary. Moreover, since the beginning of the COVID19 pandemic, the interest in TMPRSS2 has focused only on its role as a serine protease involved in the activation of the SARS-CoV-2 spike protein. However, as a soluble protease, TMPRSS2 may have additional substrates and in vitro studies have demonstrated that PAR2 is one of these substrates ^32, 33^. PAR2 is expressed in several tissues, including lung, vascular endothelial and vascular smooth muscle cells ^34,35^ and its protease-mediated activation promotes inflammation by inducing prostaglandin synthesis and cytokine production in the lungs and other organs ^36,37,38,39,40^. An intriguing hypothesis is that, similar to other soluble serine proteases, such as the human airway trypsin-like protease HAT (also known as TMPRSS11D), the soluble wild type TMPRSS2 protease may also have a role in promoting inflammation in the lungs and other tissues.

In conclusion, the T allele of the common *TMPRSS2* variant rs12329760 confers a reduced risk of severe COVID19. Similar to what observed in the TMPRSS2 KO mouse, the Val160Met substitution, which exerts a partial inhibitory effect on the proteolytic autocleavage of TMPRSS2 and the priming of the SARS-CoV-2 spike protein, is associated with a milder COVID19 infection compared to the wild type. Differences in population frequency of this genetic variant may partially contribute to the reported variability in COVID19 severity across various ethnicities. Further studies are needed to assess the expression of the TMPRSS2 across different age groups; indeed a reduced TMPRSS2 expression in young compared to older individuals, as observed in mice and in preliminary human studies, could help explain age-related differences in COVID19 morbidity. Moreover, TMPRSS2 could be a viable drug target in COVID19 patients, and camostat mesilate (a drug used to treat chronic pancreatitis and postoperative reflux esophagitis), or other novel TMPRSS2 inhibitors may have a role in the treatment of COVID19. Clinical trials are needed to confirm this prediction.

## Methods

### TMPRSS2 three-dimensional structure and variant analysis

A 3D structural model of the TMPRSS2 protein was generated using our in-house Phyre homology modelling algorithm ^41^. The FASTA sequence of TMPRSS2 was obtained from the UniProt protein knowledge database ^42^ (UniProt Id O15393, corresponding to 492 amino acid transcript Ensembl ID ENST00000332149.10). We deposited the 3D coordinates of the model in our PhyreRisk ^43^ database (http://phyrerisk.bc.ic.ac.uk/search?action=fresh-search&searchTerm=TMPRSS2). For completeness, SWISS-MODEL^44^ was also implemented to model TMPRSS2.

Model quality was assessed using: i. VoroMQA (Voronoi tessellation-based Model Quality Assessment) ^45^, a statistical tool based on inter-atomic and solvent contact areas, which returns both a local and global score between 0 and 1. A global score > 0.4 indicates a good model and < 0.3 a poor model; models with >0.3 score <0.4 cannot be correctly classified; and ii. ProSA ^46^ which returns: 1. the Z-score, which indicates the overall model quality of the target structure compared to the Z-score for all experimentally determined structures in PDB (X-rays and NMR); 2. the residue energy, which indicates the local model quality and is calculated over a 10- and 40-residue window. Negative energy values indicate good quality structure.

Given the similarity of the Phyre and the SWISS-MODEL 3D models assessed using the root mean square deviation (RMSD) (Phyre versus SWISS-MODEL model=0.30Å RMSD), all subsequent analyses should not be sensitive to which model is selected. The set of 3D coordinates from the Phyre model (VoroMQA score=0.462, and ProSA Z score=-8.96, coordinates available for download at http://phyrerisk.bc.ic.ac.uk/search?action=fresh-search&searchTerm=TMPRSS2) was used to for the structural analysis of variants.

The impact of each variant on the TMPRSS2 protein structure was assessed by analysing the following 16 features, using our in house algorithm Missense3D ^47^: breakage of a disulfide bond, hydrogen bond or salt bridge, introduction of a buried proline, clash, introduction of hydrophilic residue, introduction of a buried charged residue, charge switch in a buried residue, alteration in secondary structure, replacement of a charged with uncharged buried residue, introduction of a disallowed phi/psi region, replacement of a buried glycine with any other residue, alteration in a cavity, replacement of cis proline, buried to exposed residue switch, replacement of a glycine located in a bend. In addition, we used the SIFT ^48^ and Polyphen2 ^49^ variant predictors, which mainly use evolutionary conservation to assess the effect of each variant.

The effect of variant rs12329760 was further assess using: i. CONDEL ^50^, which reports a weighted average of the scores from fatHMM and MutationAssessor, and ii. FoldX5 force field ^51^, which calculates the stability of a protein based on the estimation of its free energy. A ΔΔG> 0.5 kcal/mol (calculated as: ΔΔG= ΔG_wt_ - ΔG_mut_) was predicted to have a destabilizing effect.

### Participants

*Genetics Of Mortality In Critical Care (GenOMICC) and the International Severe Acute Respiratory Infection Consortium (ISARIC) Coronavirus Clinical Characterisation Consortium (4C) (ISARIC 4C)*

#### Cases

this cohort comprises of 2,244 cases who passed quality control: 2,109 hospitalized, critically-ill, COVID19 positive patients admitted to 208 UK intensive care units (ICUs) and recruited as part of the GenOMICC project, and 135 severe, hospitalized COVID19 positive cases recruited as part of the International Severe Acute Respiratory Infection Consortium (ISARIC) Coronavirus Clinical Characterisation Consortium (4C) study. Only unrelated individuals (up to 3^rd^ degree, based on kinship analysis (King 2.1)) were included. Samples were excluded if the genotype-based sex inference did not match reported sex, or if XXY karyotype was present.

#### Controls

ancestry-matched controls (ratio 1 case to 5 controls) without a positive COVID19 test were obtained from the UK BioBank population study. Only unrelated individuals (up to 3^rd^ degree) were included. Individuals with sex mismatch were excluded. For validation, 45,875 unrelated individuals of European ancestry from the 100,000 Genomes Project were used as an alternative control group.

DNA extraction, genotyping and quality control have been described in detail previously ^3^. Genetic ancestry was inferred using ADMIXTURE and reference individuals from the 1000 Genomes project. Imputation was performed using the TOPMed reference panel.

### Cells, pseudovirus and plasmid

Human embryonic kidney 293T cells (293Ts; ATCC) were maintained in Dulbecco’s modified Eagle’s medium (DMEM), 10% foetal calf serum (FCS), 1% non-essential amino acids (NEAA), 1% penicillin-streptomycin (P/S) at 37°C, 5% CO_2_.

Lentiviral pseudotype production was performed as previously described ^20,31^. ACE2 FLAG was used as previously described ^20^ and TMPRSS2 expression plasmid was a kind gift from Roger Reeves (Addgene plasmid #53887; http://n2t.net/addgene:53887; RRID:Addgene_53887 ^52^. Non-cleavable ACE2-FLAG and TMPRSS2 mutants were generated by overlap extension PCR or site-directed mutagenesis.

### Phenotypic assays

293Ts were co-transfected with FLAG-tagged, non-cleavable ACE2 and TMPRSS2 as previously described ^20^. Briefly, confluent 10cm^2^ dishes of 293T cells were co-transfected with 1µg each of TMPRSS2 and ACE2-FLAG. 24 hours later, cells were resuspended in fresh media and either spun down for lysis and western blot or added to 96 well plates along with pseudovirus. 24 hours later, media was refreshed and a further 24 hours later, cells were lysed with reporter lysis buffer (Promega) and luminescence (measured as relative luminescence units, RLU) was read on a FLUOstar Omega plate reader (BMF Labtech) using the Luciferase Assay System (Promega).

Cell pellets for western blot were lysed in RIPA buffer (150mM NaCl, 1% NP-40, 0.5% sodium deoxycholate, 0.1% SDS, 50mM TRIS, pH 7.4) supplemented with an EDTA-free protease inhibitor cocktail tablet (Roche). Cell lysates were combined with 4x Laemmli buffer (Bio-Rad) with 10% β-mercaptoethanol and boiled for 5 minutes. Membranes were probed with mouse anti-tubulin (abcam; ab7291), rabbit anti-TMPRSS2 (abcam; ab92323) and/or mouse anti-FLAG (F1804, Sigma). Near infra-red (NIR) secondary antibodies, IRDye® 680RD Goat anti-mouse (abcam; ab216776) and IRDye® 800CW Goat anti-rabbit (abcam; ab216773) were subsequently used. Blots were imaged using the Odyssey Imaging System (LI-COR Biosciences). Densitometry was performed using ImageJ.

### Statistical analysis

The association between the TMPRSS2 rs12329760 variant and COVID19 severity was assessed using logistic regression. Genetic associations in the GenOMICC/ISARIC 4C cohort were analysed as previously described ^7^. Briefly, logistic regression with additive and recessive models was performed in PLINKv1.9, adjusting for sex, age, mean-centered age-squared, top 10 principal components (principal component analysis [PCA] performed to adjust for population stratification) and deprivation index decile based on UK postcode. Each major ancestry group alternative in the 100,000 Genomes control group was performed with mixed model association tests in SAIGE ^53^ (v0.39), including age, sex, age-squared, age-sex interaction and the first 20 principal components as covariates. Trans-ethnic meta-analysis of GenOMICC data for different ancestries was performed by METAL using an inverse-variance weighted method and the P-value for heterogeneity was calculated with Cochran’s Q-test for heterogeneity implemented in the same software ^54^.

Data are presented as mean±standard deviation. Log-normality was assessed using the Shapiro-Wilk test and QQ plot. A two-tailed Student’s t-test was used to compare the means of two groups. One-way ANOVA was used to compare the means of more than two groups.

## Data Availability

Full summary-level data in support of the findings of this study are available for download from [https://genomicc.org/data](https://genomicc.org/data). Individual level data can be analysed by qualified researchers in the ISARIC 4C/GenOMICC data analysis platform by application at [https://genomicc.org/data] (https://genomicc.org/data). BioBank data and Genomics England data are available to registered researchers at https://www.ukbiobank.ac.uk/ and https://www.genomicsengland.co.uk/. The COVID19 Host Genetics Initiative2 (COVID19hg) summary statistics are available at https://www.covid19hg.org/.

## Acknowledgement

This research was conducted using the UK BioBank Resource under project 788

## Ethics

Research ethics committees (Scotland 15/SS/0110, England, Wales and Northern Ireland: 19/WM/0247). Current and previous versions of the study protocol are available at genomicc.org/protocol. All participants gave informed consent.

## Conflict of inter est statement

On behalf of all authors, the corresponding author states that there is no conflict of interest

## Author contributions

NP, EP-C and JKB contributed to population data analysis. AD, TK and MJES contributed to 3D modelling and structural analysis. TPP and WSB contributed to laboratory work. NP, EP-C, TPP and AD contributed to data analysis. NP, TPP, WSB, AD contributed to study design. NP, TPP, EP-C, AC, VS-S, J-LC, LA, WSB, JKB, MJES and AD contributed to interpretation of findings and manuscript preparation. AD conceived the study, contributed to study coordination and wrote the first draft of the manuscript. All authors approved the final version of the manuscript.

